# Psychosocial Impact on Parents of Children with Strabismus: A Study of Demographic and Clinical Correlates

**DOI:** 10.1101/2025.02.28.25323110

**Authors:** Juan Ding, Wei Wang, Wei Zhang, Yueping Li

## Abstract

**Purpose:** This study aimed to explore the psychosocial challenges experienced by parents of children with strabismus and to identify demographic and clinical factors associated with these challenges.

**Methods:** A cross-sectional study was conducted involving 220 parents of children diagnosed with strabismus. Demographic and clinical characteristics of the children were collected and analyzed. The Hospital Anxiety and Depression Scale (HADS) and the Zung Self-Rating Anxiety/Depression Scale (SAS/SDS) were administered to assess parental anxiety and depression levels.

**Results:** Significant differences in anxiety and depression levels were observed between the HADS and SAS/SDS assessments. Specifically, the prevalence of anxiety was higher using HADS-A compared to SAS (21.82% vs. 12.73%, P = 0.012), while depression rates were higher using SDS compared to HADS-D (31.82% vs. 16.82%, P < 0.001). HADS-D scores were significantly associated with the severity of strabismus, parental education level, and family residence, whereas SDS scores only correlated with parental education. No significant associations were found between HADS-A/SAS scores and patient characteristics. Additionally, the HADS assessment required significantly less time to complete than the SAS/SDS (2.54 ± 1.45 minutes vs. 7.25 ± 4.13 minutes, P < 0.001).

**Conclusion:** Identifying psychosocial distress among parents of children with strabismus is crucial for comprehensive care. The HADS emerges as a more efficient and effective tool for assessing emotional well-being compared to the SAS/SDS, highlighting its potential utility in clinical and research settings.

**Highlights:**

- Study examines psychological issues in parents of children with strabismus.
- HADS and SAS/SDS used to assess anxiety and depression in 220 parents.
- HADS-A vs. SAS and HADS-D vs. SDS show significant difference in detection rates.
- HADS-D score correlates with strabismus severity, parental education, and residence.
- HADS is more time-efficient than SAS/SDS, making it suitable for clinical settings.

## Introduction

Strabismus, characterized by misaligned visual axes, is a prevalent eye condition that typically emerges during critical periods of visual development^[1]^. Occurring in 0.01% to 7% of the population, strabismus is not only an individual matter but a public health concern^[2–7]^. Beyond the physical implications such as amblyopia and impaired binocular vision, it can also lead to significant psychosocial challenges including diminished self-esteem, tension in interpersonal relationships, and reduced employment opportunities^[9–13]^ .

Previous research has predominantly concentrated on the intensity of patients’ emotional responses from a psychopathological perspective ^[14]^. Tools like the Health-Related Quality of Life (HRQOL) and the Hospital Anxiety and Depression Scale (HADS) have been crafted to evaluate the emotional state of patients receiving medical care for physical ailments^[15]^. The impact of intermittent exotropia (IXT) on children’s mental health can be further scrutinized using proxy scales^[16]^. Studies have shown that strabismus that begins in childhood has long-lasting effects, deteriorating the HRQOL for both the affected children and their parents.^[17–19]^. Moreover, recent research has highlighted that the emotional state of parents significantly influences the psychosocial well-being of their children with strabismus. Successful surgical intervention for strabismus can markedly enhance the psychosocial function and life quality of the child and the family as a whole^[20]^. It is therefore essential to diagnose and confirm anxiety and/or depressive symptoms in parents, as this plays a pivotal role in the subsequent treatment and well-being of the children.

Instruments such as the Zung Self-Rating Anxiety/Depression Scale (SAS/SDS) and HADS are widely utilized across various medical disciplines including dermatology, psychiatry, rheumatology, orthopedics, nephrology, and oncology^[14,21–25]^.However, there is a dearth of research that delves into and compares the efficacy of HADS and SAS/SDS in evaluating the emotional well-being of parents with strabismic children. This study aims to fill that gap by assessing anxiety and depression in parents of Chinese children with strabismus using both HADS and SAS/SDS scales. Additionally, it seeks to determine the correlation between the scores obtained from these scales and the clinical indicators of the children’s condition, thereby providing valuable insights for future clinical research into the psychological aspects of parents with strabismic children.

## Method

### Participants

The aims of the study were meticulously explained to each participant. The study was conducted among parents of children under 14 years of age with strabismus, in the inpatient department of , who were scheduled for their children’s strabismus surgeries the following day. Exclusion criteria were established to ensure participants’ physical and mental well-being did not impede their ability to accurately respond to the study’s assessment scales.

Data were collected on demographic characteristics, clinical data, and psychosocial measures. The clinical data include the type of strabismus, prior ocular history, binocular visual function, and previous history of strabismus surgery. Prism cover test, and an alternative cover test were used to determine deviation and angle of strabismus.

All processes were carried out in accordance with the applicable norms and legislation. Ethics approval for this research was granted by the Medical Ethics Committee of (#2022050). Consent from participants was collected in March 22, 2022, and all subjects provided consent to share their information for research purposes.

Regarding the access to identifying information, authors had access to information that could identify individual participants during data collection for the purpose of accurately recording and analyzing the data. After data collection, the identifying information was securely stored and only accessible by authorized researchers who needed it for the study.

### Instruments

HADS (Hospital Anxiety and Depression Scale). A 14-item scale is divided into anxiety and depression subscales of 7 items each (even for depression and odd for anxiety). Each item is graded on a Likert scale of 0 to 3, with higher scores indicating more severe anxiety and despair. The combined HADS-A and HADS-D scores ranged from 0 to 21.HADS anxiety was defined as a HADS-A score ≥ 8, and HADS depression was defined as a HADS-D score ≥ 8. Anxiety and depression severity was further graded as follows: (1) mild, 8-10; (2) moderate,11-14; and (3) severe ,15-21 ^[26]^ .

The combined Zung SAS and SDS scores ranged from 0 to 100, and the Zung SAS score ≥50 was considered to indicate SAS anxiety, while the Zung SDS score ≥ 50 indicated SAD depression. Gradings for the intensity of anxiety and depression were mild (50-59), moderate (60-69), and severe (70-100) ^[27,28]^. After thoroughly learning the contents of the scales, the participants would independently complete the HADS and SAS/SDS self-reports. The HADS and SAS/SDS scores would then be calculated and recorded by the nurses who were blind to the design of the study.

## Results

### Patient and parents’ general characteristics

A total of 220 parents of children with strabismus engaged in this study. All were adults, with ages ranging from 24 to 62 years, averaging 36.07± 6.04 years. Females constituted a significant majority of the participants, at 77.72% (171 out of 220). The majority of the families hailed from urban areas, representing 66.36% (146 out of 220), in contrast to their rural counterparts, which accounted for 33.63% (74 out of 220).

Regarding educational attainment, the participants’ qualifications were diverse: 47.27% (104 out of 220) held bachelor’s degrees, making it the most common level of education among them. This was followed by high school diplomas at 40.91% (90 out of 220), master’s degrees at 6.63% (14 out of 220), and primary education at 5.45% (12 out of 220). In terms of understanding their children’s condition, a sizable majority, 63.18% (139 out of 220), admitted to not fully understanding their child’s strabismus. Conversely, 35.91% (79 out of 220) demonstrated a comprehensive understanding of strabismus, while a very small percentage, 0.91% (2 out of 220), were completely unaware.Table 1 provides a detailed summary of the clinical characteristics of the children with strabismus.

**Table 1.**
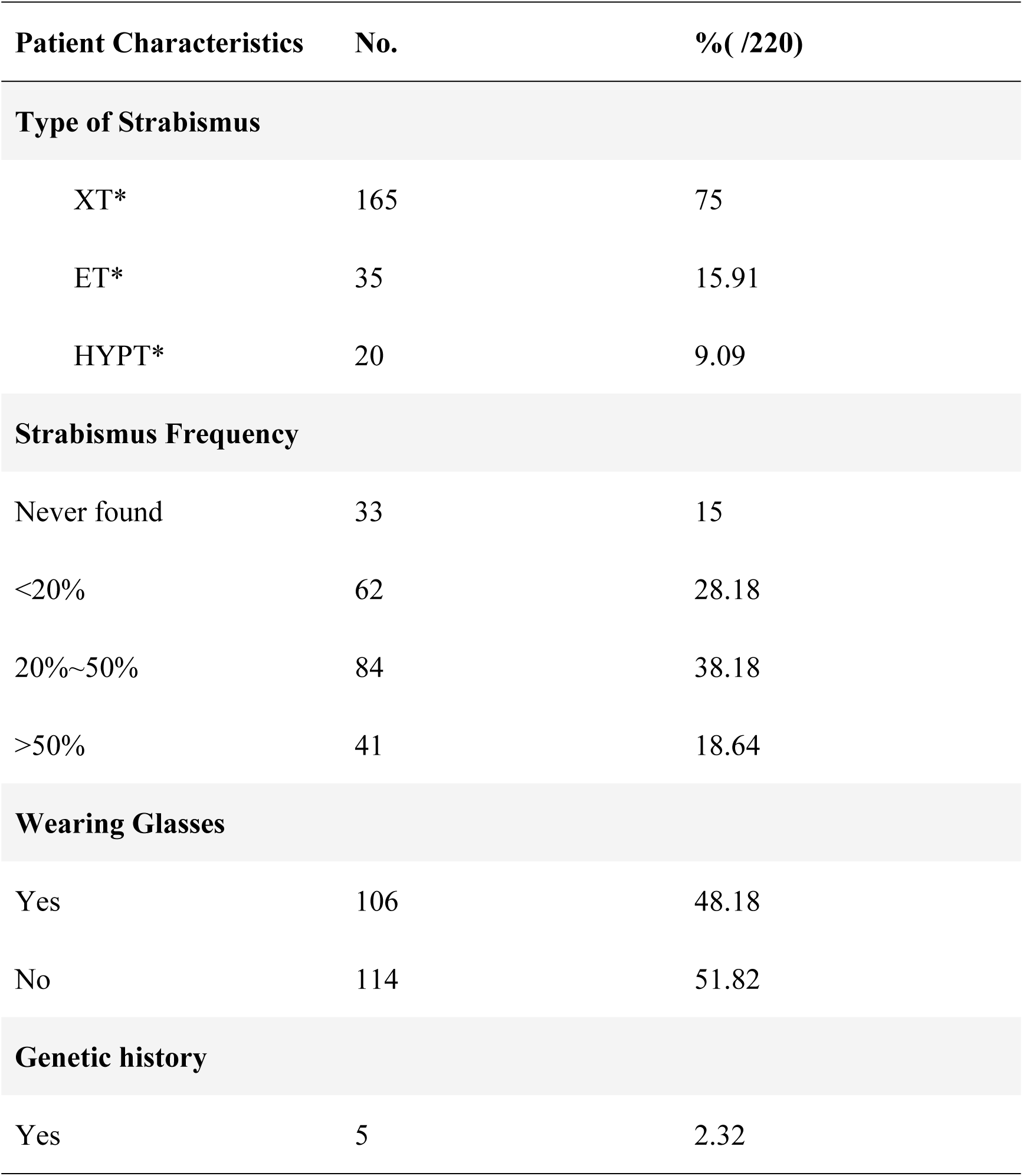
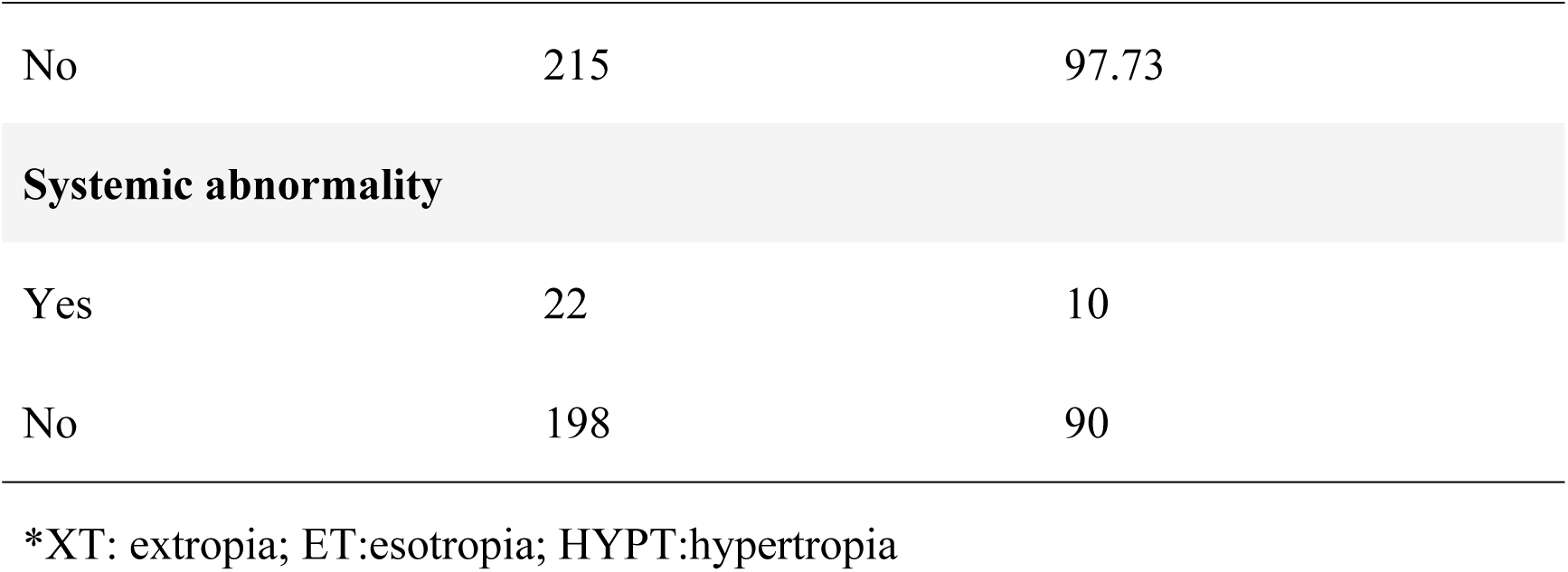
Clinical features of strabismic patients.

| Patient Characteristics | No. | %( /220) |
| --- | --- | --- |
| **Type of Strabismus** |  |  |
| XT* | 165 | 75 |
| ET* | 35 | 15.91 |
| HYPT* | 20 | 9.09 |
| **Strabismus Frequency** |  |  |
| Never found | 33 | 15 |
| <20% | 62 | 28.18 |
| 20%~50% | 84 | 38.18 |
| >50% | 41 | 18.64 |
| **Wearing Glasses** |  |  |
| Yes | 106 | 48.18 |
| No | 114 | 51.82 |
| **Genetic history** |  |  |
| Yes | 5 | 2.32 |
| No | 215 | 97.73 |
| **Systemic abnormality** |  |  |
| Yes | 22 | 10 |
| No | 198 | 90 |

\*XT: extropia; ET:esotropia; HYPT:hypertropia

### Comparison of Anxiety Detection in Parents of Children with Strabismus between HADS-A and SAS

The study discerned a marked discrepancy in the prevalence of anxiety among parents of children with strabismus when evaluated using the HADS-A and SAS scales, with 21.82% and 12.73% of parents, respectively, identified as anxious (P = 0.012, as shown in Fig. 1a). An in-depth analysis of the data revealed that the majority of parents with concurrent anxiety were categorized as mildly anxious, with 77.09% identified by HADS-A and 92.86% by SAS. Moderate anxiety was detected in 20.83% of parents by HADS-A and 7.14% by SAS, while severe anxiety affected 2.08% as per HADS-A, and none by SAS (Fig. 1b). Notably, the severity of anxiety, as evaluated by both the HADS-A and SAS criteria, did not reveal a statistically significant difference (Fisher test, *P* = 0.192).

**Fig. 1.**
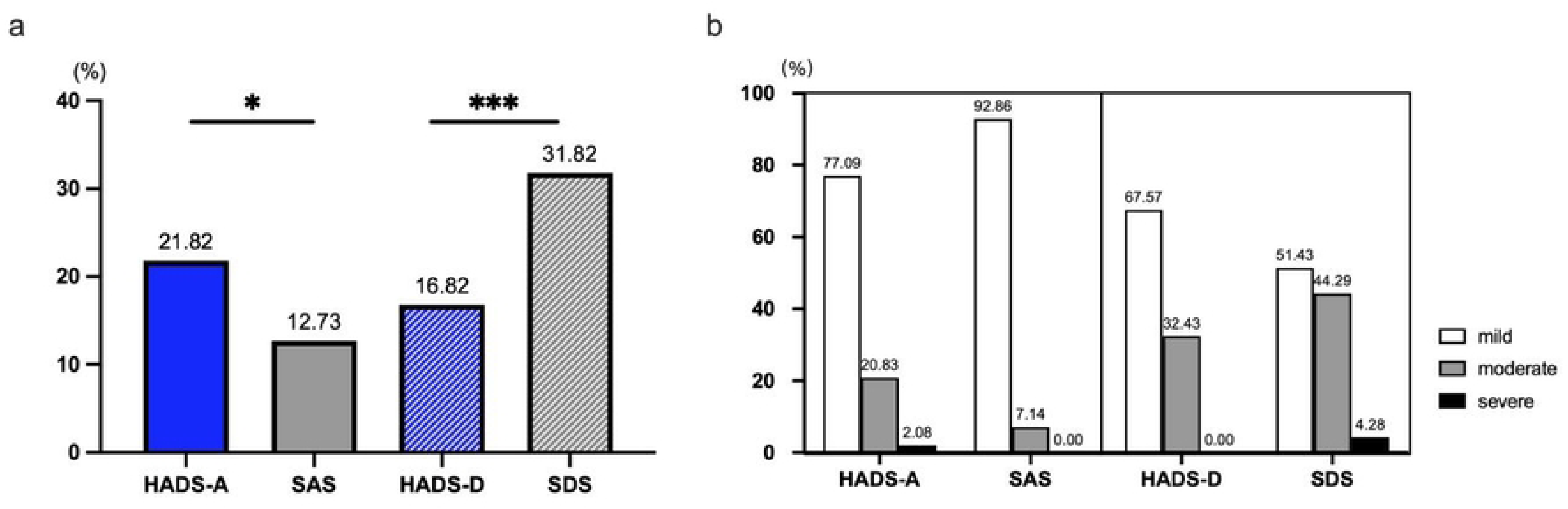
Detection rate and severity of anxiety and depression in parents of children with strabismus by HADS-A and SAS, HADS-D and SDS. (a) Significant difference in detection rates between HADS-A and SAS, HADS-D and SDS was noted. (b) No discernible variation in the percentage of severity between HADS-A and SAS, HADS-D and SDS .\**P*<0.05,\**P*<0.01

Further insights from Figure 2 indicate that a small proportion, 5.91% (13 out of 220), of parents met the criteria for anxiety based on both HADS-A and SAS. Anxiety was uniquely identified by HADS-A in 15.91% (35 out of 220) of parents and solely by SAS in 6.82% (15 out of 220). The remaining majority, 71.36% (157 out of 220), did not manifest symptoms of anxiety according to either set of criteria. A significant correlation was observed between the scores obtained from HADS-A and SAS (Spearman’s rank correlation test, r = 0.468, P < 0.001, Fig.2), highlighting a strong positive relationship between the two methods of anxiety assessment.

**Fig. 2.**
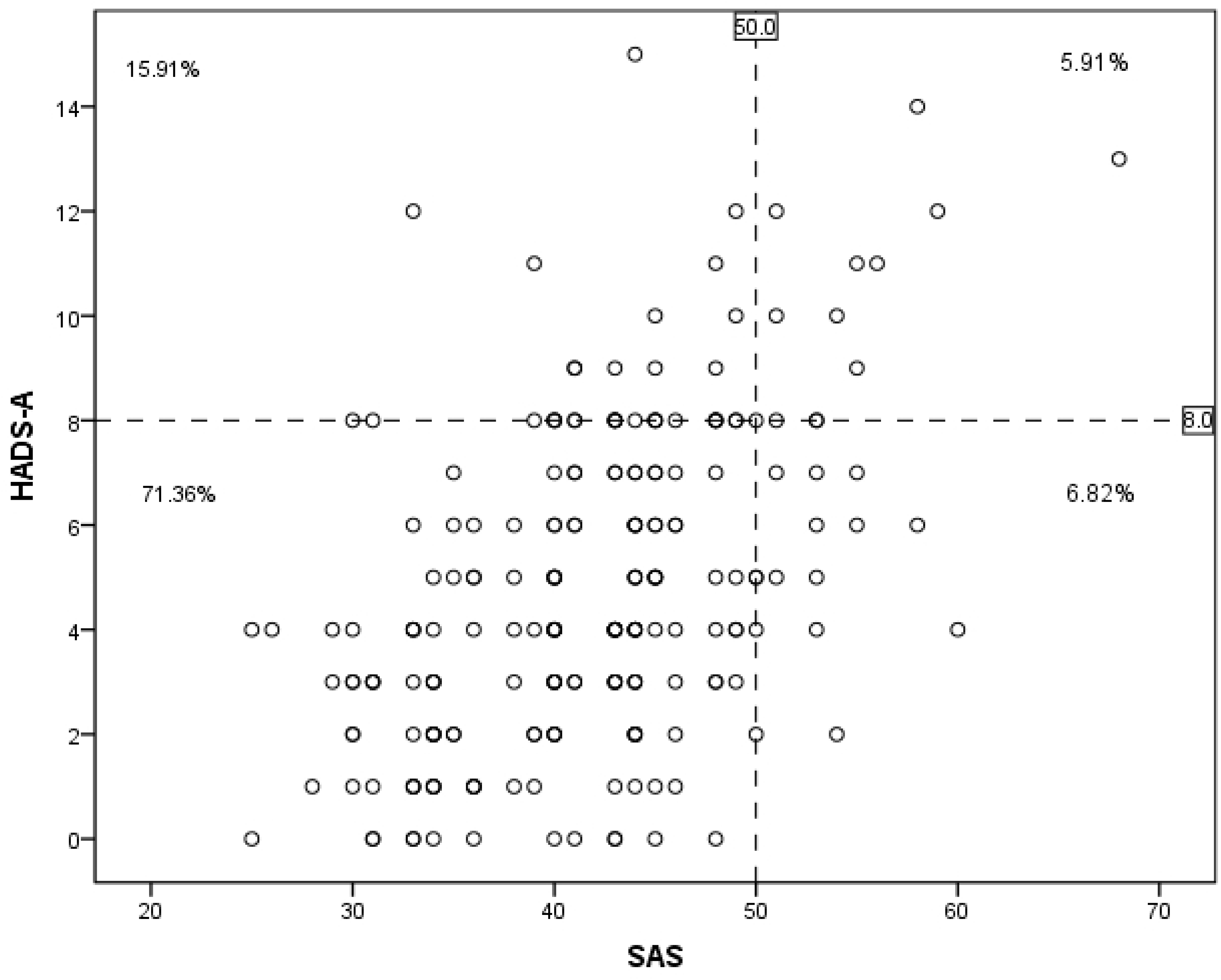
Correlation of HADS-A and SAS in parents of children with strabismus. Positive correlation between the score of HADS-A and SAS was observed.

### Comparison of Depression Detection in Parents of Children with Strabismus between HADS-D and SDS

Figure 1a delineates a significant difference in the detection of depression among parents of children with strabismus when assessed by the HADS-D and SDS scales, with rates of 16.82% and 31.82%, respectively (*P* < 0.001). A comparative analysis of the depression severity according to HADS-D and SDS criteria revealed the following distribution among affected parents: mild depression was identified in 67.57% by HADS-D and 51.43% by SDS, moderate depression in 32.43% by HADS-D and 44.29% by SDS, and severe depression in none by HADS-D and 4.28% by SDS (as depicted in Figure 1b). The assessment of depression severity through these criteria showed no significant difference (Fisher test, *P* = 0.190).

As seen in Figure 3, 10.91% (24/220) of the patients met the criteria for depression according to both HADS-D and SDS, 5.91% (13/220) met the criteria for depression according to only HADS-D, 20.91% (46/220) met the criteria for depression according to only SDS, and 62.27% (137/220) did not show anxiety according to either HADS-A or SAS. There was a significant correlation between the HADS-D and SDS scores(Spearman test, r = 0.523, *P* = 0.000). (Fig 3)

**Fig.3.**
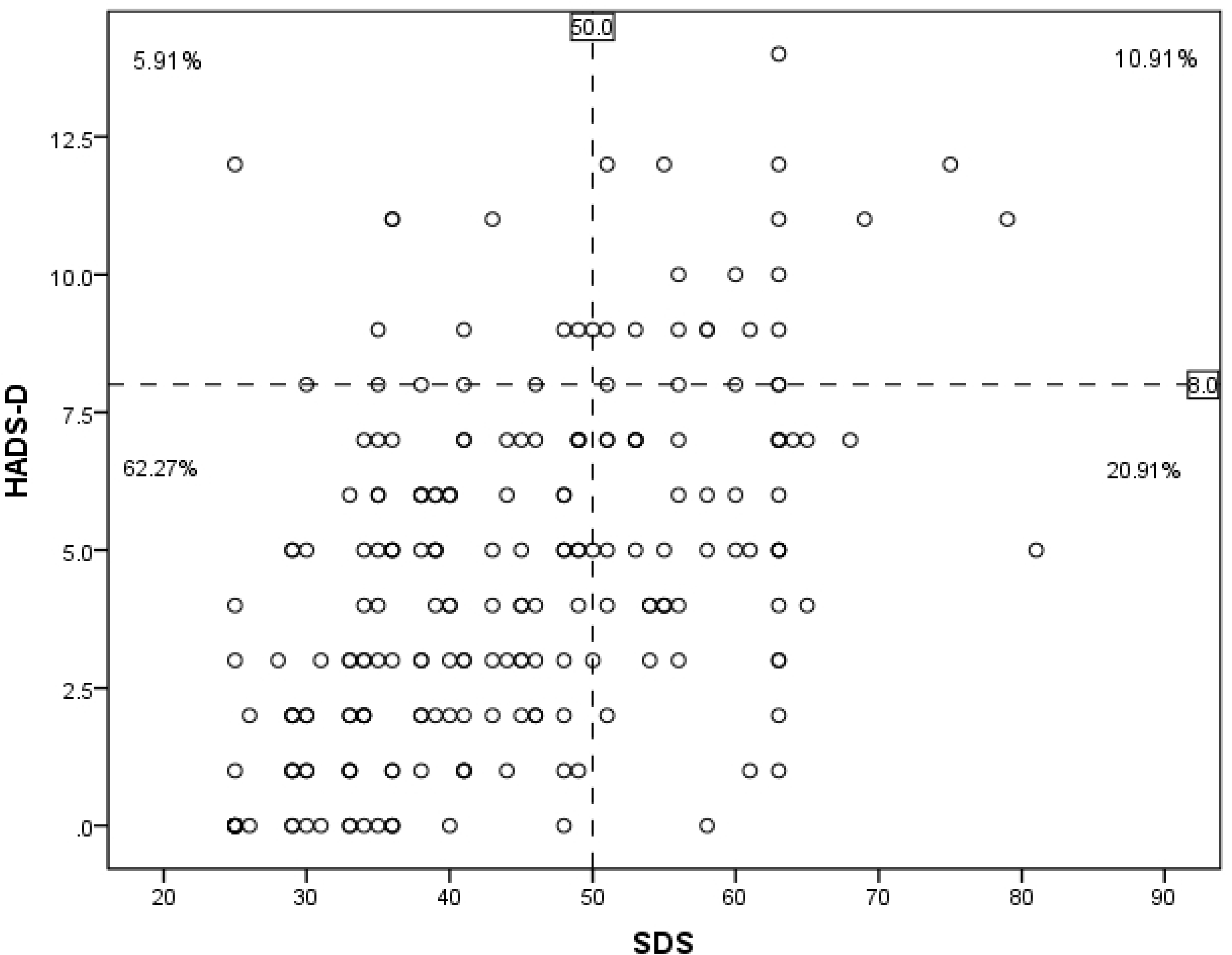
Correlation of SDS and HADS-D in parents of children with strabismus. A significant correlation was found between SDS and HADS-D.

### Correlation between the clinical data of patients and the anxiety and depression scores assessed by the HADS-A and SAS, HADS-D and SDS in parents

Tables 2 and 3 of the study do not indicate any significant correlation between the SAS scores and HADS-A scores in relation to the clinical data.

**Table 2.** Correlation of anxiety and depression score with clinical data (continuous variables)

|  | HADS-A score | SAS score | HADS-D score | SDS score |
| --- | --- | --- | --- | --- |
| **Parents age** |  |  |  |  |
| Relative coefficient | 0.013 | -0.017 | -0.005 | -0.007 |
| *P* value | 0.851 | 0.802 | 0.942 | 0.914 |
| **Patients age** |  |  |  |  |
| Relative coefficient | -0.029 | -0.001 | 0.065 | -0.005 |
| *P* value | 0.667 | 0.983 | 0.340 | 0.940 |
| **Duration of strabismus** |  |  |  |  |
| Relative coefficient | 0.080 | 0.066 | 0.102 | 0.099 |
| *P* value | 0.238 | 0.332 | 0.132 | 0.143 |
| **Deviation** |  |  |  |  |
| Relative coefficient | 0.109 | 0.076 | 0.159 | 0.051 |
| *P* value | 0.106 | 0.259 | 0.018* | 0.455 |

Correlation between the variables was found using a Spearman rank correlation. $*P$ less than 0.05 was regarded as significant.

In addition to this, the study examined the relationship between the HADS-D and SDS scores and various clinical details. The HADS-D score demonstrated a significant positive correlation with the degree of strabismus deviation (*P* = 0.018, as per Table 2) and the family’s inhabitation (*P* = 0.034). Concurrently, both HADS-D and SDS scores showed a positive correlation with the level of education of the parents (*P* = 0.001 for HADS-D and *P* = 0.040 for SDS, as detailed in Table 3).

**Table 3.**
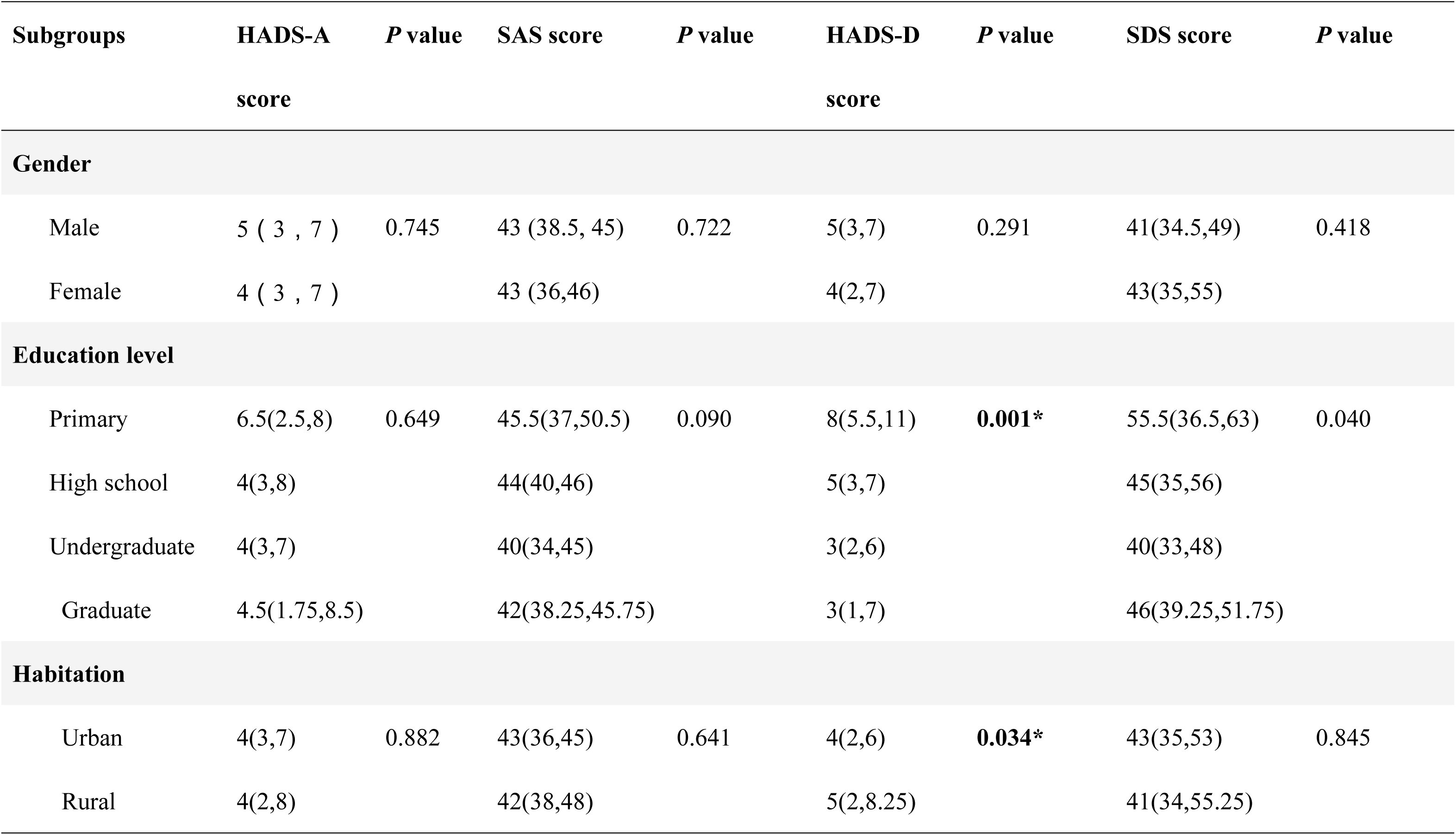
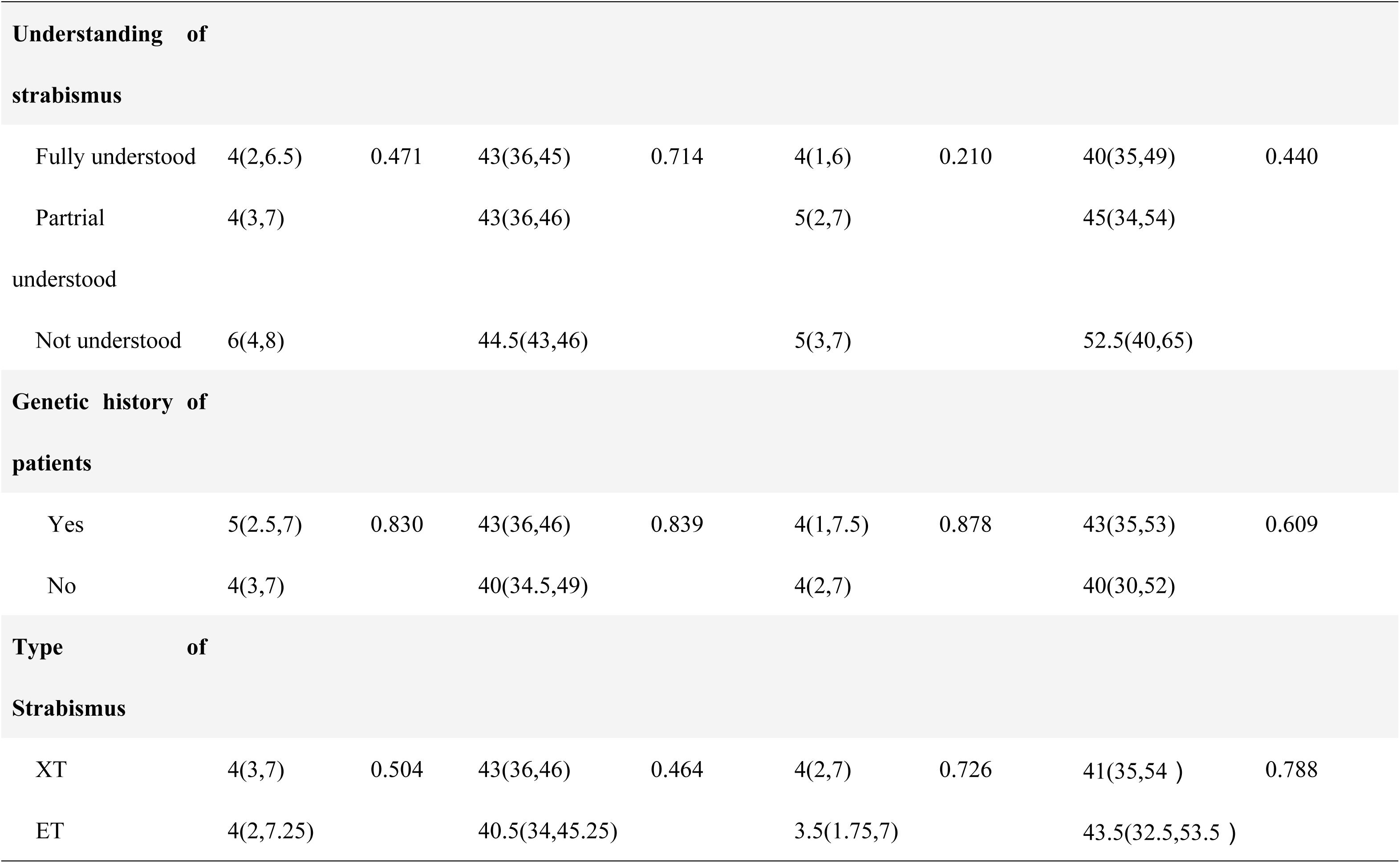
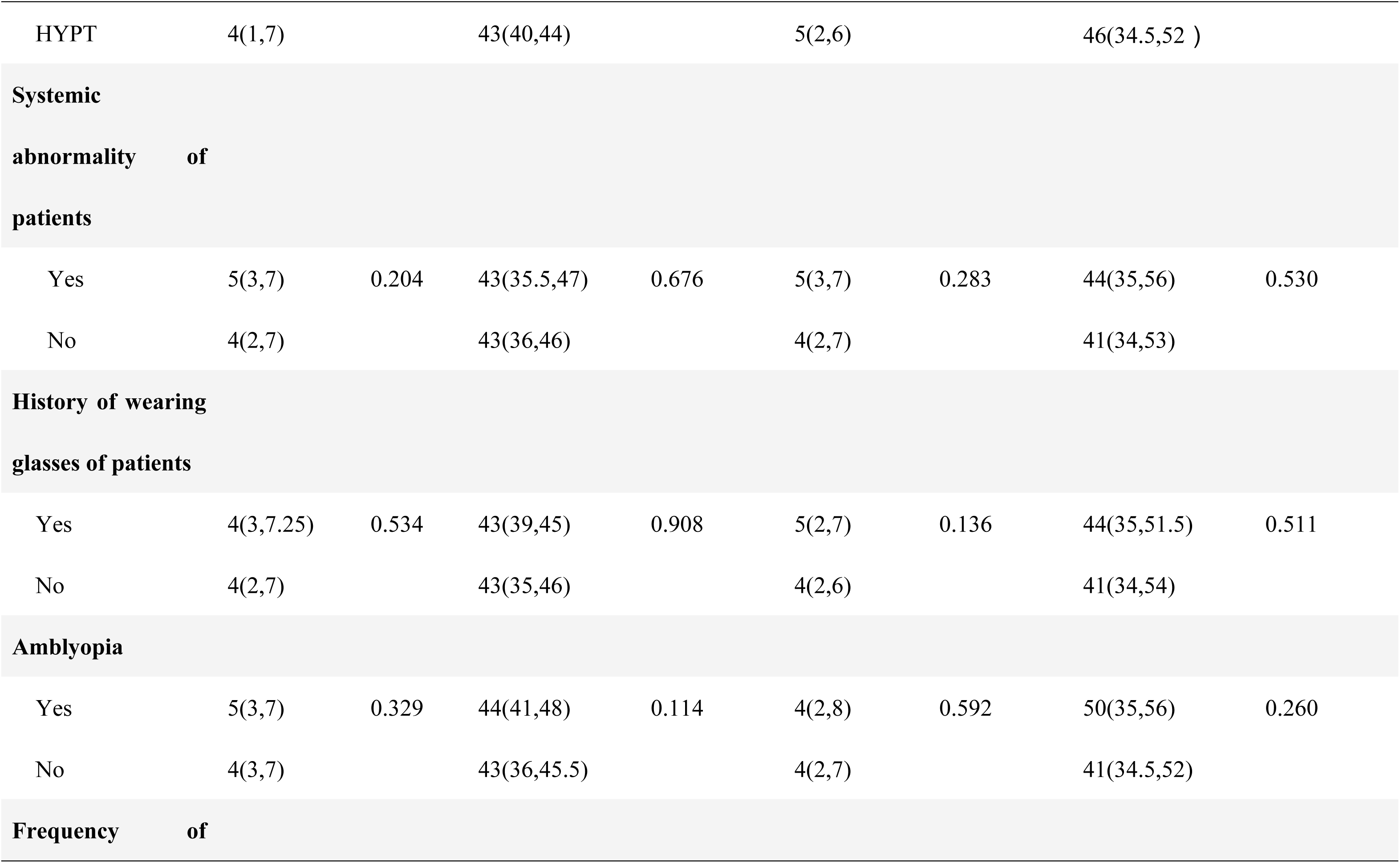
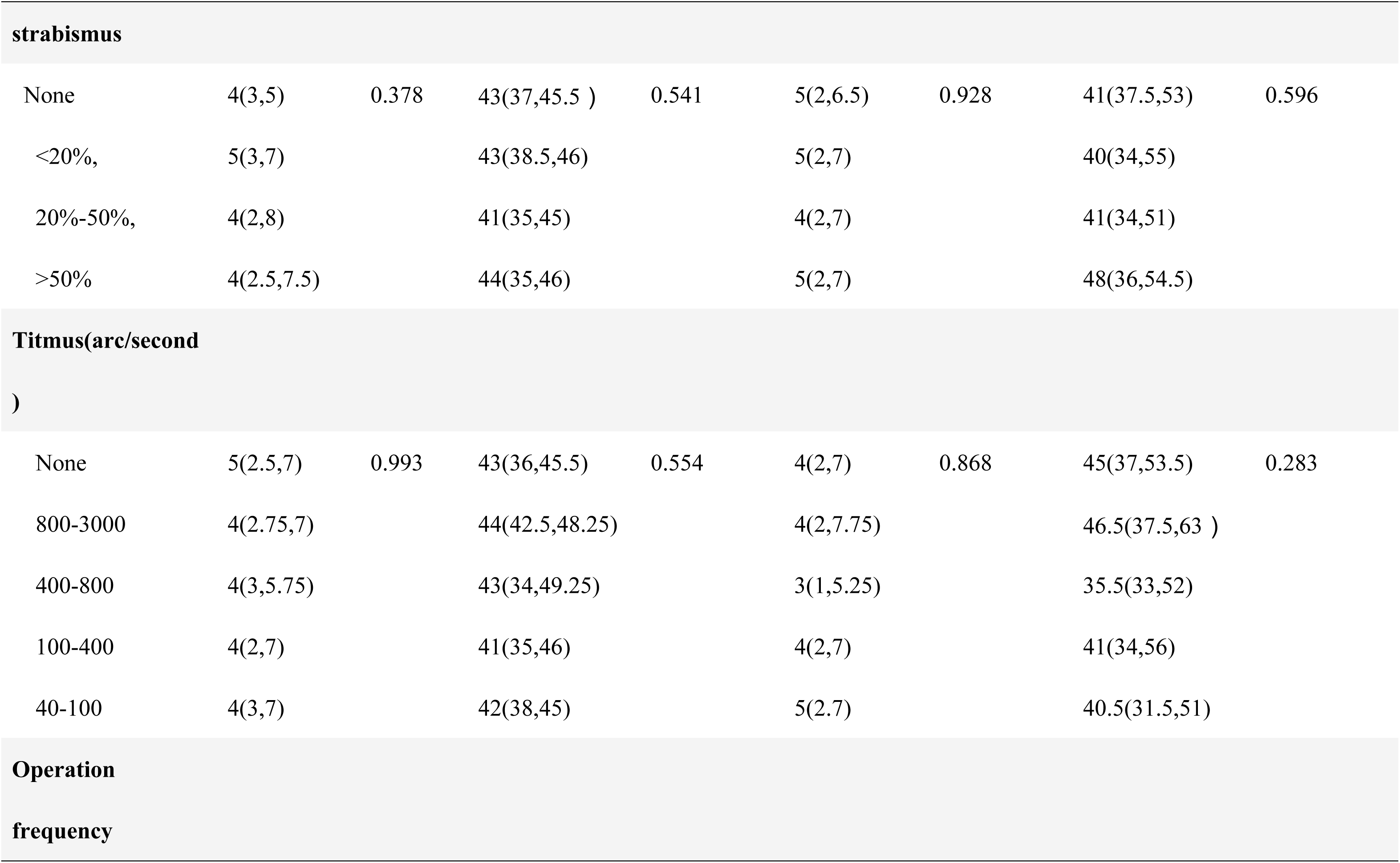
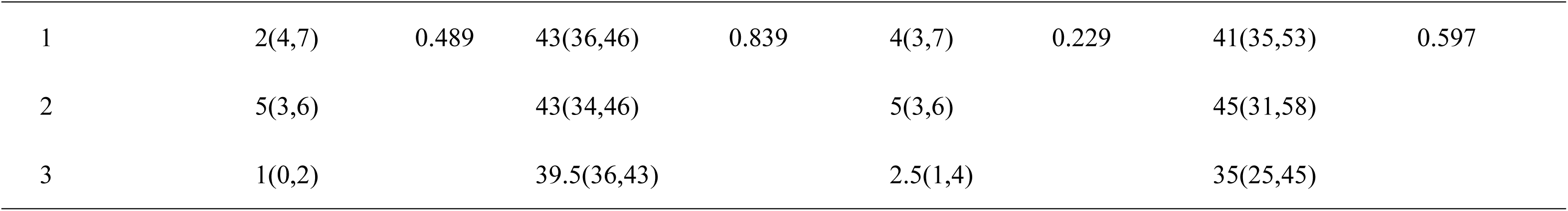
Correlation of anxiety and depression score with clinical data (discontinuous variables)

| Subgroups | HADS-A<br>score | *P* value | SAS score | *P* value | HADS-D<br>score | *P* value | SDS score | *P* value |
| --- | --- | --- | --- | --- | --- | --- | --- | --- |
| **Gender** |  |  |  |  |  |  |  |  |
| Male | 5 ( 3 , 7 ) | 0.745 | 43 (38.5, 45) | 0.722 | 5(3,7) | 0.291 | 41(34.5,49) | 0.418 |
| Female | 4 ( 3 , 7 ) |  | 43 (36,46) |  | 4(2,7) |  | 43(35,55) |  |
| **Education level** |  |  |  |  |  |  |  |  |
| Primary | 6.5(2.5,8) | 0.649 | 45.5(37,50.5) | 0.090 | 8(5.5,11) | **0.001\*** | 55.5(36.5,63) | 0.040 |
| High school | 4(3,8) |  | 44(40,46) |  | 5(3,7) |  | 45(35,56) |  |
| Undergraduate | 4(3,7) |  | 40(34,45) |  | 3(2,6) |  | 40(33,48) |  |
| Graduate | 4.5(1.75,8.5) |  | 42(38.25,45.75) |  | 3(1,7) |  | 46(39.25,51.75) |  |
| **Habitation** |  |  |  |  |  |  |  |  |
| Urban | 4(3,7) | 0.882 | 43(36,45) | 0.641 | 4(2,6) | **0.034\*** | 43(35,53) | 0.845 |
| Rural | 4(2,8) |  | 42(38,48) |  | 5(2,8.25) |  | 41(34,55.25) |  |

| Understanding of strabismus |  |  |  |  |  |  |  |  |
| --- | --- | --- | --- | --- | --- | --- | --- | --- |
| Fully understood | 4(2,6.5) | 0.471 | 43(36,45) | 0.714 | 4(1,6) | 0.210 | 40(35,49) | 0.440 |
| Partial understood | 4(3,7) |  | 43(36,46) |  | 5(2,7) |  | 45(34,54) |  |
| Not understood | 6(4,8) |  | 44.5(43,46) |  | 5(3,7) |  | 52.5(40,65) |  |
| Genetic history of patients |  |  |  |  |  |  |  |  |
| Yes | 5(2.5,7) | 0.830 | 43(36,46) | 0.839 | 4(1,7.5) | 0.878 | 43(35,53) | 0.609 |
| No | 4(3,7) |  | 40(34.5,49) |  | 4(2,7) |  | 40(30,52) |  |
| Type of Strabismus |  |  |  |  |  |  |  |  |
| XT | 4(3,7) | 0.504 | 43(36,46) | 0.464 | 4(2,7) | 0.726 | 41(35,54 ) | 0.788 |
| ET | 4(2,7.25) |  | 40.5(34,45.25) |  | 3.5(1.75,7) |  | 43.5(32.5,53.5 ) |  |
| HYPT | 4(1,7) |  | 43(40,44) |  | 5(2,6) |  | 46(34.5,52 ) |  |
| **Systemic abnormality of patients** |  |  |  |  |  |  |  |  |
| Yes | 5(3,7) | 0.204 | 43(35.5,47) | 0.676 | 5(3,7) | 0.283 | 44(35,56) | 0.530 |
| No | 4(2,7) |  | 43(36,46) |  | 4(2,7) |  | 41(34,53) |  |
| **History of wearing glasses of patients** |  |  |  |  |  |  |  |  |
| Yes | 4(3,7.25) | 0.534 | 43(39,45) | 0.908 | 5(2,7) | 0.136 | 44(35,51.5) | 0.511 |
| No | 4(2,7) |  | 43(35,46) |  | 4(2,6) |  | 41(34,54) |  |
| **Amblyopia** |  |  |  |  |  |  |  |  |
| Yes | 5(3,7) | 0.329 | 44(41,48) | 0.114 | 4(2,8) | 0.592 | 50(35,56) | 0.260 |
| No | 4(3,7) |  | 43(36,45.5) |  | 4(2,7) |  | 41(34.5,52) |  |
| **Frequency of** |  |  |  |  |  |  |  |  |

| strabismus |  |  |  |  |  |  |  |  |
| --- | --- | --- | --- | --- | --- | --- | --- | --- |
| None | 4(3,5) | 0.378 | 43(37,45.5 ) | 0.541 | 5(2,6.5) | 0.928 | 41(37.5,53) | 0.596 |
| <20%, | 5(3,7) |  | 43(38.5,46) |  | 5(2,7) |  | 40(34,55) |  |
| 20%-50%, | 4(2,8) |  | 41(35,45) |  | 4(2,7) |  | 41(34,51) |  |
| >50% | 4(2.5,7.5) |  | 44(35,46) |  | 5(2,7) |  | 48(36,54.5) |  |
| Titmus(arc/second ) |  |  |  |  |  |  |  |  |
| None | 5(2.5,7) | 0.993 | 43(36,45.5) | 0.554 | 4(2,7) | 0.868 | 45(37,53.5) | 0.283 |
| 800-3000 | 4(2.75,7) |  | 44(42.5,48.25) |  | 4(2,7.75) |  | 46.5(37.5,63 ) |  |
| 400-800 | 4(3,5.75) |  | 43(34,49.25) |  | 3(1,5.25) |  | 35.5(33,52) |  |
| 100-400 | 4(2,7) |  | 41(35,46) |  | 4(2,7) |  | 41(34,56) |  |
| 40-100 | 4(3,7) |  | 42(38,45) |  | 5(2,7) |  | 40.5(31.5,51) |  |
| Operation frequency |  |  |  |  |  |  |  |  |
| 1 | 2(4,7) | 0.489 | 43(36,46) | 0.839 | 4(3,7) | 0.229 | 41(35,53) | 0.597 |
| 2 | 5(3,6) |  | 43(34,46) |  | 5(3,6) |  | 45(31,58) |  |
| 3 | 1(0,2) |  | 39.5(36,43) |  | 2.5(1,4) |  | 35(25,45) |  |

Data is presented as the median with interquartile ranges (25th to 75th percentiles). For comparing two independent groups, the Mann-Whitney U test was employed, while the Kruskal-Wallis H rank-sum test was utilized for the analysis of three or more independent groups. The study considered \**P* < 0.05 and \**P* < 0.01 as thresholds for statistical significance.

### Time consumption between HADS and Zung SAS/SDS

HADS took 2.54±1.45 minutes on average, which was less than Zung SAS/SDS (7.25±4.13minutes) (*P*<.0001) (Fig. 4).

**Fig.4.**
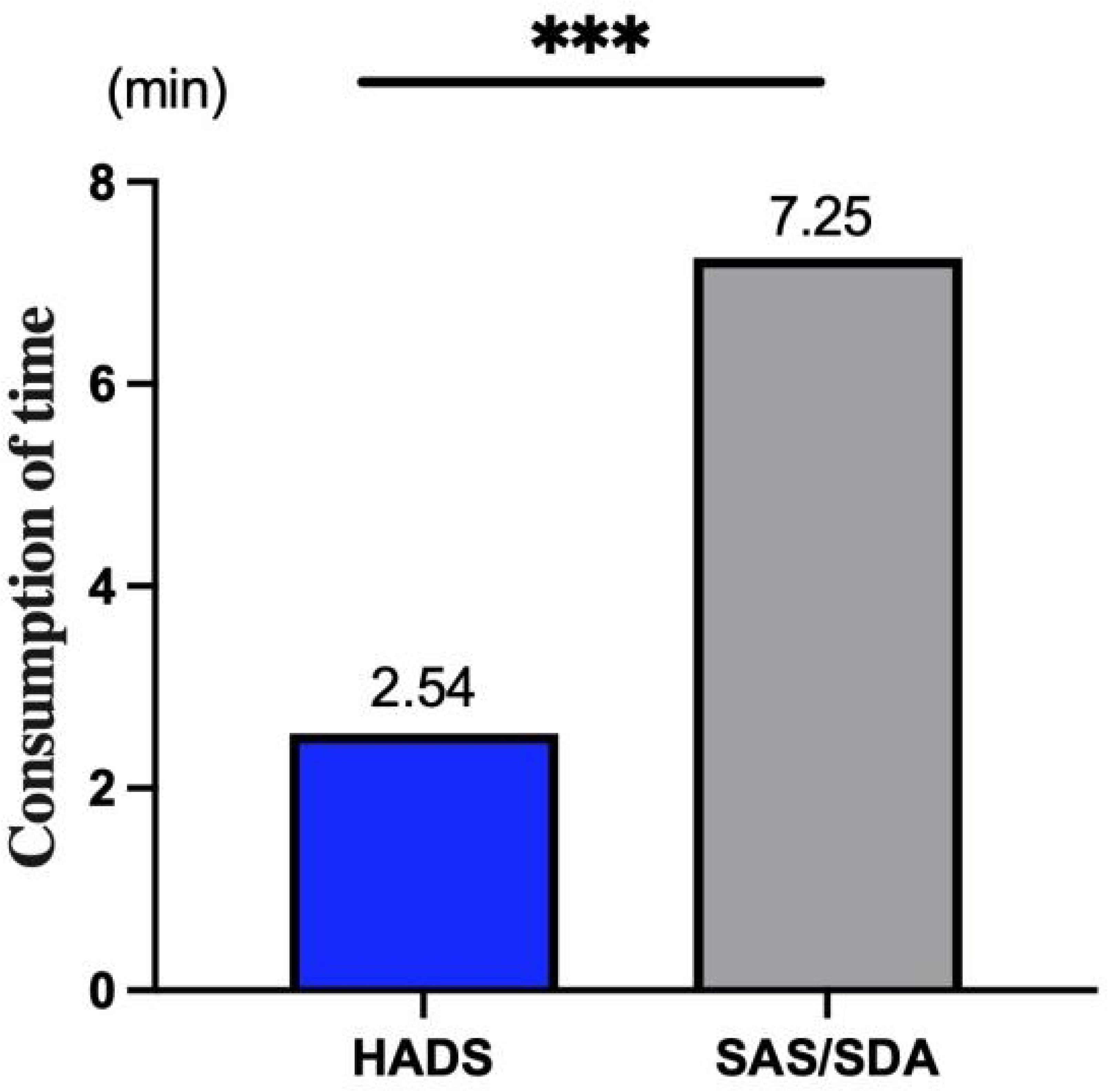
HADS and SAS/SDS assessment time spent by parents of children with strabismus. Compared to SAS/SDS, HADS required less time for assessment. * P < 0.01 was considered significant.

## Discussion

In recent years, the psychological state of children with strabismus and their families has been followed with interest increasingly. According to a series of studies, parents of children who have intermittent exotropia (IXT) show a clear predisposition toward psychological suffering based on their anxiety and depressive symptoms. Since more than 90% of IXT patients exhibit a decline in their mental health, the parents are genuinely worried about their children ^[20]^.Encouragingly, successful strabismus surgery has been shown to have a markedly positive effect on the psychosocial functioning of both the children and their families, notably improving anxiety levels and overall quality of life.

In clinical practice, psychological assessment by a professional remains the gold standard for diagnosing anxiety and depression. Nonetheless, self-report measures such as HADS and SAS/SDS are frequently utilized for their efficiency in screening and quantifying these conditions. The HADS, a self-administered questionnaire, is designed to assess the impact of mood disorders, with a focus on anxiety and depression, on overall medical patient distress. Initially created for hospital use, the HADS has since gained broader application, extending to community and family health screenings, including those for patients with various medical conditions ^[29]^.The SAS/SDS, another valuable tool in this domain, is employed to gauge the intensity of a patient’s anxiety or depressive symptoms over the preceding week. These 20-item scales provide a quantitative assessment that aids in identifying the severity of affective disturbances, offering valuable insights for further clinical evaluation and intervention.

According to a prior study, based on HADS criteria, anxiety and depression are diagnosed in 24% and 11% of adult strabismus patients, respectively ^[30]^. These figures are intriguingly in line with our findings concerning parents of children with strabismus. In our study, the positive rates of anxiety and depression among these parents were 21.82% and 16.82%, respectively—approximately an order of magnitude higher than the rates observed in the general population ^[31]^. The anxiety rate by HADS-A was higher than that by SAS, and the depression detection rate by HADS-D was lower than that of SDS in our study, based on the HADS and SAS/SDS criteria. The higher detection of anxiety by HADS-A may be attributed to its sensitivity to immediate stressors, such as a child’s hospitalization, unlike the SAS, which evaluates the psychological state over a more recent timeframe.We postulate that the SDS, with its broader range of questions and potential to identify more cases of depression, might contribute to the perceived heightened risk of depressive symptoms.

Despite notable differences in the detection rates of anxiety and depression between the HADS-A and SAS, and between the HADS-D and SDS, the severity of these conditions as measured by both scales remained consistent.Correlation analysis showed that HADS-A and SAS had a strong agreement in identifying anxiety, as did HADS-D and SDS in identifying depression. In our study, clinical levels of anxiety and depression were reported by 5.91% (13/220) and 10.91% (24/220) of parents, respectively. Had these individuals been assessed by a mental health professional, they might have received a psychiatric diagnosis.

Additionally, HADS exhibited a more robust correlation with clinical data of the patients than the SAS/SDS. Our findings indicated a significant positive correlation between the HADS-D scores and the degree of strabismus deviation, as well as factors such as parental education level and whether the family resided in urban or rural settings. As the angle of ocular misalignment increases, the visibility of the condition becomes more pronounced, potentially heightening parental concerns and exacerbating symptoms of psychological distress, including depression. This trend is consistent with previous studies that have reported similar findings ^[32, 33]^.Moreover, parents of children with strabismus who were diagnosed with depression were often found to have lower levels of education and to reside in rural areas. We hypothesize that the interplay between limited education and scarce medical resources in rural China may amplify the visibility of the condition and intensify the lack of social support and understanding of strabismus among these families. These factors could contribute to the increased psychological burden experienced by affected parents.

Furthermore, the administration of the HADS was more time-efficient compared to the SAS/SDS, aligning with previous findings [34]. This efficiency can be attributed to the brevity of the HADS questionnaire, its focused topics, and straightforward scoring system. In the case of chronic conditions like strabismus, which necessitate multiple follow-up consultations before and after surgery, a swift assessment tool is particularly beneficial. The expedited nature of the HADS can minimize the time commitment for follow-up visits, potentially increasing the willingness of family members to engage in the process. Given these advantages, we suggest that HADS may be a more suitable and acceptable tool for clinical settings.

The study faced certain limitations: firstly, being a single-center clinical study, it was susceptible to selection bias due to regional constraints; secondly, its cross-sectional design lacked follow-up, hindering the evaluation of the long-term effectiveness of the HADS and SAS/SDS scales in assessing anxiety and depression.

## Conclusion

To enhance the success of strabismus surgery, it is crucial to identify parents of children with strabismus who are undergoing considerable psychosocial distress and to gain insights into their expectations following the surgery. In the quest to assess anxiety and depression within this particular demographic of parents, the Hospital Anxiety and Depression Scale (HADS) presents itself as a practical and effective measure.

### Relevance for clinical practice

The findings underscore the significant and lasting impact that childhood-onset strabismus can have on a patient’s health-related quality of life. This study brings to light the fact that parents of children with strabismus are also subjected to considerable psychosocial distress, an issue that necessitates recognition and intervention. The Hospital Anxiety and Depression Scale (HADS) is put forward as a viable and potent tool for evaluating the levels of anxiety and depression among parents of children afflicted with strabismus.

## Data Availability

All relevant data are within the manuscript and its Supporting Information files.

## Declaration

### Ethics approval and consent to participate

All methods were performed by following per under the relevant guidelines and regulations. Ethics approval for this research was granted by the Medical Ethics Committee of the Tianjin Eye Hospital(#2022050). Informed consent was collected and all subjects provided consent to share their information for research purposes.

### Authors’ contributions

Dr. YPL has full access to the data and takes overall responsibility. Conception and design: YPL, JD; Data collection and collation: WW, JD, WZ; Data analysis and interpretation: JD, YPL; Writing: JD. All authors read and approved the final manuscript.

## Acknowledgments

We are deeply grateful to all participants who made contributions to our study for their generous participation.

## Data availability statement

The data that support the findings of this study are available from the corresponding author upon reasonable request.

